# Genomic analysis of SARS-CoV-2 breakthrough infections from Varanasi, India

**DOI:** 10.1101/2021.09.19.21262487

**Authors:** Lamuk Zaveri, Royana Singh, Priyoneel Basu, Sofia Banu, Payel Mukherjee, Shani Vishwakarma, Chetan Sahni, Manpreet Kaur, Nitish Kumar Singh, Abhay Kumar Yadav, Ajay Kumar Yadav, Ashish, Shivani Mishra, Shivam Tiwari, Surendra Pratap Mishra, Amareshwar Vodapalli, Himasri Bollu, Debashruti Das, Prajjval Pratap Singh, Gyaneshwer Chaubey, Divya Tej Sowpati, Karthik Bharadwaj Tallapaka

## Abstract

Studies worldwide have shown that the available vaccines are highly effective against SARS-CoV-2. However, there are growing laboratory reports that the newer variants of concerns (VOCs e.g. Alpha, Beta, Delta etc) may evade vaccine induced defense. In addition to that, there are few ground reports on health workers having breakthrough infections. In order to understand VOC driven breakthrough infection we investigated 14 individuals who tested positive for SARS-CoV-2 after being administered a single or double dose of Covishield (ChAdOx1, Serum Institute of India) from the city of Varanasi, which is located in the Indian state of Uttar Pradesh. Genomic analysis revealed that 78.6% (11/14) of the patients were infected with the B.1.617.2 (Delta) variant. Notably, the frequency (37%) of this variant in the region was significantly lower (p<0.01), suggesting that the vaccinated people were asymmetrically infected with the Delta variant. Most of the patients tested displayed mild symptoms, indicating that even a single dose of the vaccine can help in reducing the severity of the disease. However, more comprehensive epidemiological studies are required to understand the effectiveness of vaccines against the newer VOCs.

## Introduction

With the growing knowledge on SARS-CoV-2 spread and infection, it is now clear that accomplishing community immunity by natural infections is more putative than ground reality. Therefore, in the global fight against SARS-CoV-2, vaccinating the entire population against the virus is an important step in slowing down the pandemic. By establishing community immunity with the help of vaccines, it might be possible to prevent further large outbreaks. The Government of India initiated the vaccination programme against SARS-CoV-2 in January 2021 with the aim of vaccinating the entire Indian population by the end of 2021. In the initial phase, the Government targeted frontline workers at highest risk to the disease. With greater availability of vaccines, the scope of vaccination was widened to include people above the age of 65 and then above 45. Due to the staggered nature of vaccination and the onset of a large second wave, reports emerged of people testing positive for SARS-CoV-2 despite being vaccinated [1–4]. At a total population of approximately 1 billion, the time taken to vaccinate the entire Indian population would be quite long. This is further complicated by the fact that most SARS-CoV-2 vaccines approved for use are double dose vaccines with a minimum interval of four weeks between the two doses [1]. Partial immunity in the population has led to a situation where there are variants of SARS-CoV-2 emerging against which existing vaccines have decreased efficacy [6].

Reports of breakthrough infections have impacted the public confidence in the efficacy of these vaccines. However, a majority of these infections are in healthcare workers who have repeated and high exposure to SARS-CoV-2 unlike the population at large. Also the routine nasal testing of fully vaccinated people have shown low titers of SARS-CoV-2 [7]. Therefore, it is essential to study breakthrough infections and their outcomes among general populations. In our survey in Varanasi, a city in the Indian state of Uttar Pradesh, we found 14 individuals who tested positive for SARS-CoV-2 after receiving either a single or double dose of Covishield (ChAdOx1) vaccine. We studied in detail the virus variants, the main symptoms and the fate of these individuals due to this infection.

## Materials and Methods

### Ethical approval

This study was approved by the local institutional ethical committee. Written consent was taken from patients wherever applicable.

### Sample collection

Nasopharyngeal swab samples were collected from routine testing done at Multidisciplinary Research Unit, Banaras Hindu University, Varanasi, Uttar Pradesh, India. Samples were collected with informed consent and details on vaccination, symptoms and comorbidities were collected via a telephonic interview. RNA was extracted from a 200 μl sample using the QIAamp Viral RNA Mini Kit (Qiagen) and eluted in either 50 μl nuclease free water or RNA elution buffer provided with the kit. 20 μl of GB RNA IC (internal control) was added to lysate after the lysis to monitor quality of the preps. RT-PCR was performed for detection of COVID-19 infection using GB SARS-CoV-2 Real-Time RT-PCR kit (General Biologicals Corporation) according to manufacturer’s protocol for E gene, N gene and ORF1ab (RdRp). A SARS-CoV-2 positive control and negative control was also included in the RT-PCR to ensure accurate reporting of results.

### Whole genome sequencing

Viral RNA from patients that tested positive were shipped to CCMB, Hyderabad India. The samples were sequenced using Illumina COVIDSeq (Illumina) according to the manufacturer’s protocol. Briefly, first strand cDNA was synthesized in sets of 96 samples using random hexamers and reverse transcriptase. The viral first strand cDNA was then amplified using two primer pools. Post amplification, the two amplified pools were combined and the samples were Tagmented. The Tagmentation process fragments the PCR products and tags the fragmented products with adapter sequences. Using paramagnetic beads, the tagmented samples were purified. An amplification step adds a 10 base pair i7 index and i5 adapter required for sequencing cluster generation. The 96 samples were then pooled into a single tube and purified using paramagnetic beads. The bead purification also serves to select for optimal sized fragments required for sequencing. The purified sample was quantified and 8 such pools were normalized and then loaded onto an SP flow cell (Illumina). 384 samples were loaded per lane for a total of 768 total samples per flow cell. Reads were set for 100 bp paired end sequencing on a NovaSeq 6000 (Illumina).

### Data processing

Basecalling was performed on raw image data using bcl2fastq v2.20.0.422 (Illumina). Quality control of FASTQ files was performed using FASTQC v0.11.9 [8]. Poor quality bases and adapters were trimmed using Trimmomatic [9]. Alignment of reads to the indexed reference genome NC_045512.2 was done using HISAT2 v2.1.0 [10]. Consensus sequences were generated using bcftools from the BAM file post alignment. Coverage across the genome was calculated using samtools depth. PANGO v3.0.5 was used to assign lineages to the consensus sequences [11]. Mutations in the sequences were identified using Nextclade v1.1.0 [12]. For sequences where lineages were not designated by PANGO, a manual assignment was provided based on the presence of key lineage defining mutations.

### Phylogenetic analysis

Datasets of 444 genomes from Uttar Pradesh, and 172 global breakthrough samples deposited on GISAID (till 15th July 2020) were used for the analysis (Supplementary Table S1). The metadata and submitting institutions are listed in Supplementary Data. The sequences were aligned against the WH1 reference genome using MAFFT and IQTREE was used to construct a phylogenetic time tree using the sequence collection date [13,14]. The trees were viewed and appropriately annotated using iTOL [15].

## Results and discussion

The samples were collected in the month of April 2021 when the vaccine was available only for those above the age of 45 thus the median age of patients studied was 55. Of the 14 breakthrough samples, 4 patients were female while the remaining were males. The oldest female patient was in the 50 to 55 years age group while the oldest male patient was in the 80 to 85 years age group. The ‘youngest’ female patient was in the 50 to 55 years age group and male patient was in the 40 to 45 years age group.

The patients displayed a range of mild symptoms commonly associated with SARS-CoV-2 (Table 1) including fever and diarrhea in the initial phase of infection along with cough and body aches [16]. Only 2 patients had comorbidities, BT3 had Diabetes Type - II and BT7 had high blood pressure. None of the patients developed any severe symptoms indicating possible protective effects of the vaccine. Notably, one patient was asymptomatic and was discovered during contact tracing, highlighting the importance of this method in controlling the spread of SARS-CoV-2 [17].

**Table 1:** Patient details along with comorbidity and symptoms.

| Patient | Sex | Age | Comorbidity | Symptoms | Pango assignment |
| --- | --- | --- | --- | --- | --- |
| BT1 | Male | 51-60 | None | Fever, cough | B.1.617.2 |
| BT2 | Male | 51-60 | None | Headache, sore throat, fever | Undetermined |
| BT3 | Male | 71-80 | Diabetes Type - II | Diarrhoea, low fever | B.1.617.2 |
| BT4 | Female | 51-60 | None | Fever 3 days, neck pain | B.1.617.2 |
| BT5 | Male | 81-90 | None | Diarrhoea 3 days, mild fever 5 days | B.1.617.2 |
| BT6 | Male | 51-60 | None | Mild symptoms | B.1.617.2 |
| BT7 | Male | 51-60 | High blood pressure | No symptoms, discovered in contact tracing | B.1.617.2 |
| BT8 | Male | 51-60 | None | Mild symptoms | Undetermined |
| BT9 | Male | 41-50 | None | mild symptoms | B.1.617.2 |
| BT10 | Female | 51-60 | None | Mild symptoms | B.1.617.2 |
| BT11 | Male | 41-50 | None | Mild symptoms | Undetermined |
| BT12 | Female | 51-60 | None | Diarrhoea 3 days, fever, cough 4 days | B.1.617.2 |
| BT13 | Female | 51-60 | None | Fever, sore throat | B.1.617.2 |
| BT14 | Male | 41-50 | None | Mild symptoms | B.1.617.2 |

Of the 14 patients in this study, 10 patients tested positive after getting their first dose of the vaccine while the remaining 4 tested positive after their second dose. The median duration between the dose of the vaccine and testing positive for SARS-CoV-2 was 19 days (Figure 1). In the double dose vaccination category, the onset of the disease was 10 to 15 days post vaccination (Figure 1). Patient BT2 started showing symptoms a day post vaccination suggesting that he may have caught the disease before getting the second dose of the vaccine (Figure 1). In the single dose category, the median duration for the disease onset was 19 days from vaccination with a range of 14 to 34 days (Figure 1).

**Figure 1:**
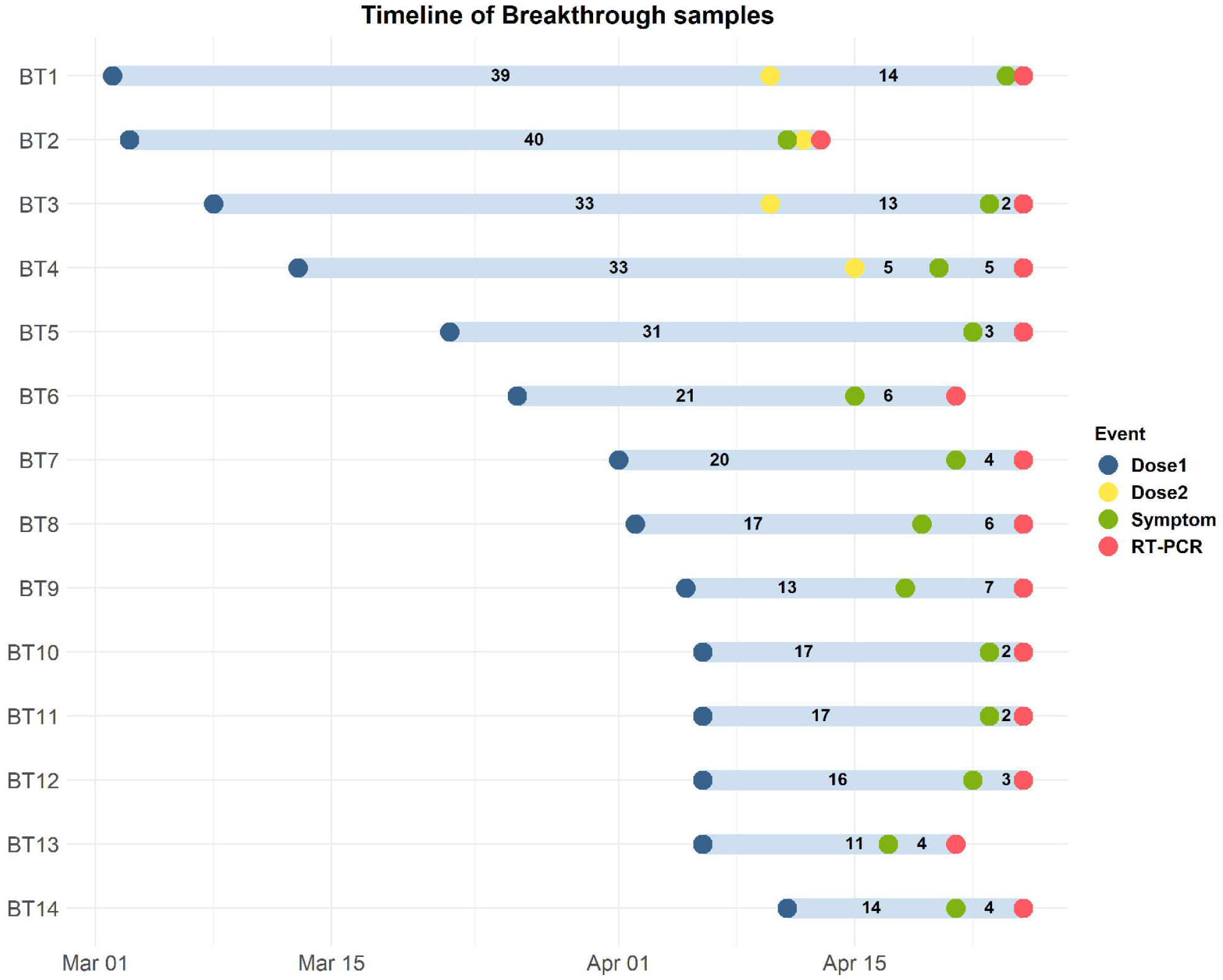
Timelines between vaccination and onset of disease. Vaccination dates, symptom onset and date of testing for each of the patients was plotted to compare the duration in days between the last vaccination dose administered, onset of the disease and confirmation of SARS-CoV-2.

Lineage assignment was done using Pango run V. 3.0.5, which revealed that 11 out of the 14 patients were infected with the B.1.617.2 (Delta) strain (Table 1). The remaining patients’ strains couldn’t be determined due to low coverage of the viral genome. Of the 4 patients who had received both doses of the vaccine, 3 were infected with the B.1.617.2 which is in line with recent studies indicating B.1.617.2 could be an immune escape variant [18,19]. From the 10 patients that received a single dose, 8 patients were infected with B.1.617.2 and the remaining could not be determined due to low coverage.

In order to understand the breakthrough infections, we compared the frequency of VOC distribution in vaccinated and unvaccinated Individuals from Varanasi. In the vaccinated individuals the frequency of the Delta variant was 0.785 (95% CI 0.488 - 0.942), whereas, in the unvaccinated, the frequency of these variants was 0.37 (95% CI 0.284 - 0.465). Considering the null model one should expect the equal distribution of VOCs among both of the groups, whilst there was a significant (Fisher’s exact test, p value <0.01) higher frequency of the Delta variant among vaccinated individuals. Thus our analysis suggests that the Delta variant can possibly have immune escape properties.

Exploring further, phylogenetic analysis of samples deposited in GISAID from Uttar Pradesh revealed the presence of multiple variants of concern including B.1.617.2 and B.1.36 (Figure 2) [20]. Analysis of samples from the month of April indicated that the dominant strains across the country were B.1.617.2 (Delta), followed by B.1.617.1 (Kappa) and B.1.1.7 (Alpha) (Figure 3). As B.1.617.2 is one of the dominant strains present in India, it is possible that Varanasi may also have it in similar proportions during that time period. However, this has to be confirmed with larger cohorts from this place.

**Figure 2:**
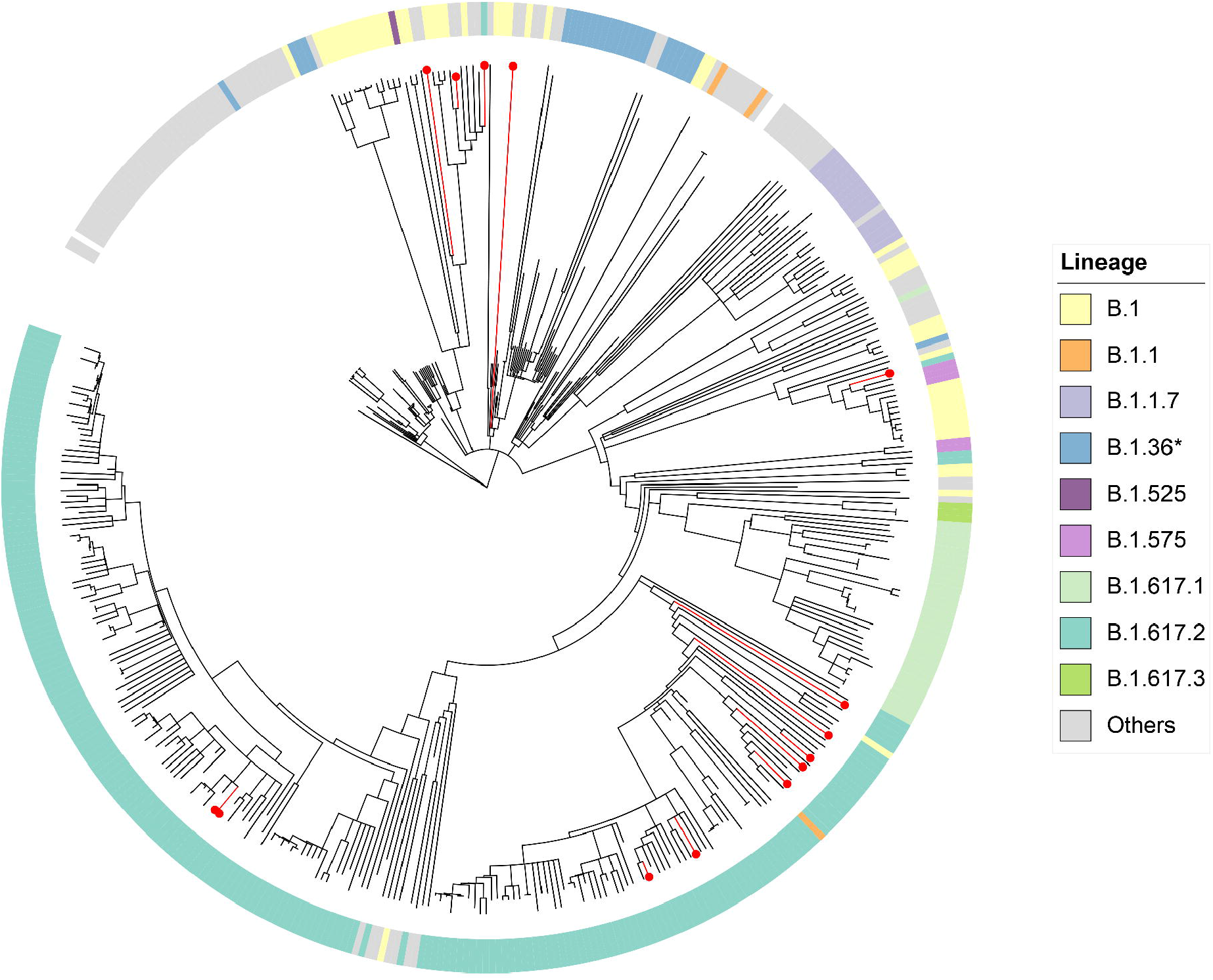
Phylogenetic analysis of samples from Uttar Pradesh, India. Phylogenetic analysis of 444 samples deposited in GISAID from Uttar Pradesh, India till April 2021. The tree shows the lineages across samples from Uttar Pradesh with the red lines indicating breakthrough samples from Varanasi. 11 of the 14 samples fall in the B.1.617.2 lineage, while the remaining 3 were not assigned a lineage due to low coverage. The dominant strain present in Uttar Pradesh is B.1.617.2 (Delta).

**Figure 3:**
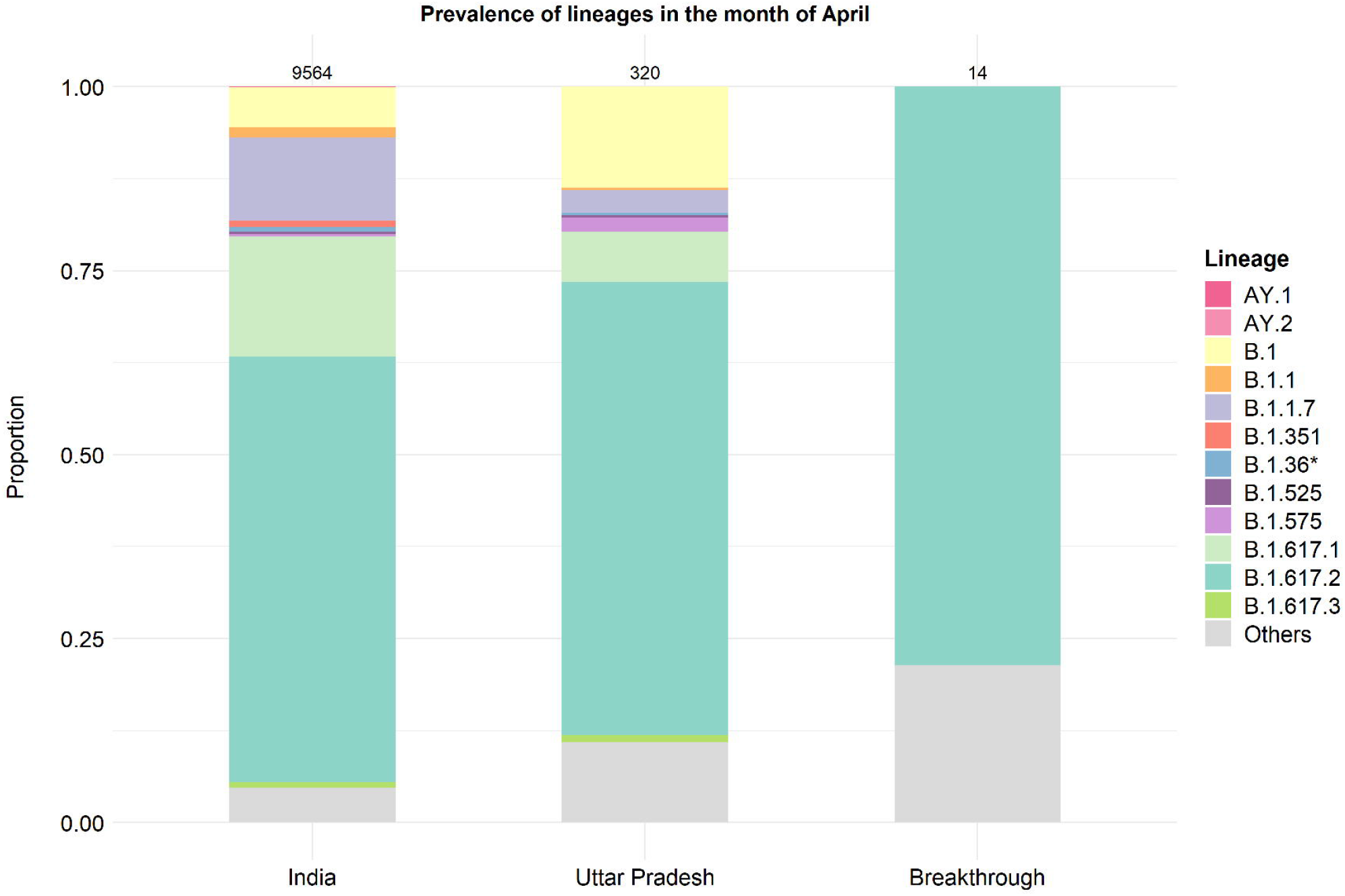
Prevalence of lineages in India and Uttar Pradesh in the month of April in context of Breakthrough infections. The plot shows Delta as the dominant variant in the month of April. Analysis of breakthrough samples revealed 11 out of 14 samples were of Delta indicating a similar trend in vaccine breakthrough infections.

Although breakthrough infections occur only in a small fraction of the population, global analysis of breakthrough samples deposited in GISAID reveals that 89% (153/172) samples belong to the current VOCs/VOIs such as B.1.1.7 (Alpha), B.1.351 (Beta), B.1.617.2 (Delta) and B.1.427/B.1.429 (Epsilon) (Figure 4). Higher proportions of breakthrough infections associated with VOCs, in light of studies revealing reduced neutralization of VOCs by vaccine sera, is a matter of concern. Achieving community immunity by rapid vaccination of populations across the world, as well as continuously monitoring the behavior of viral strains in response to existing vaccines, is imperative to prevent the evolution and spread of such VOCs.

**Figure 4:**
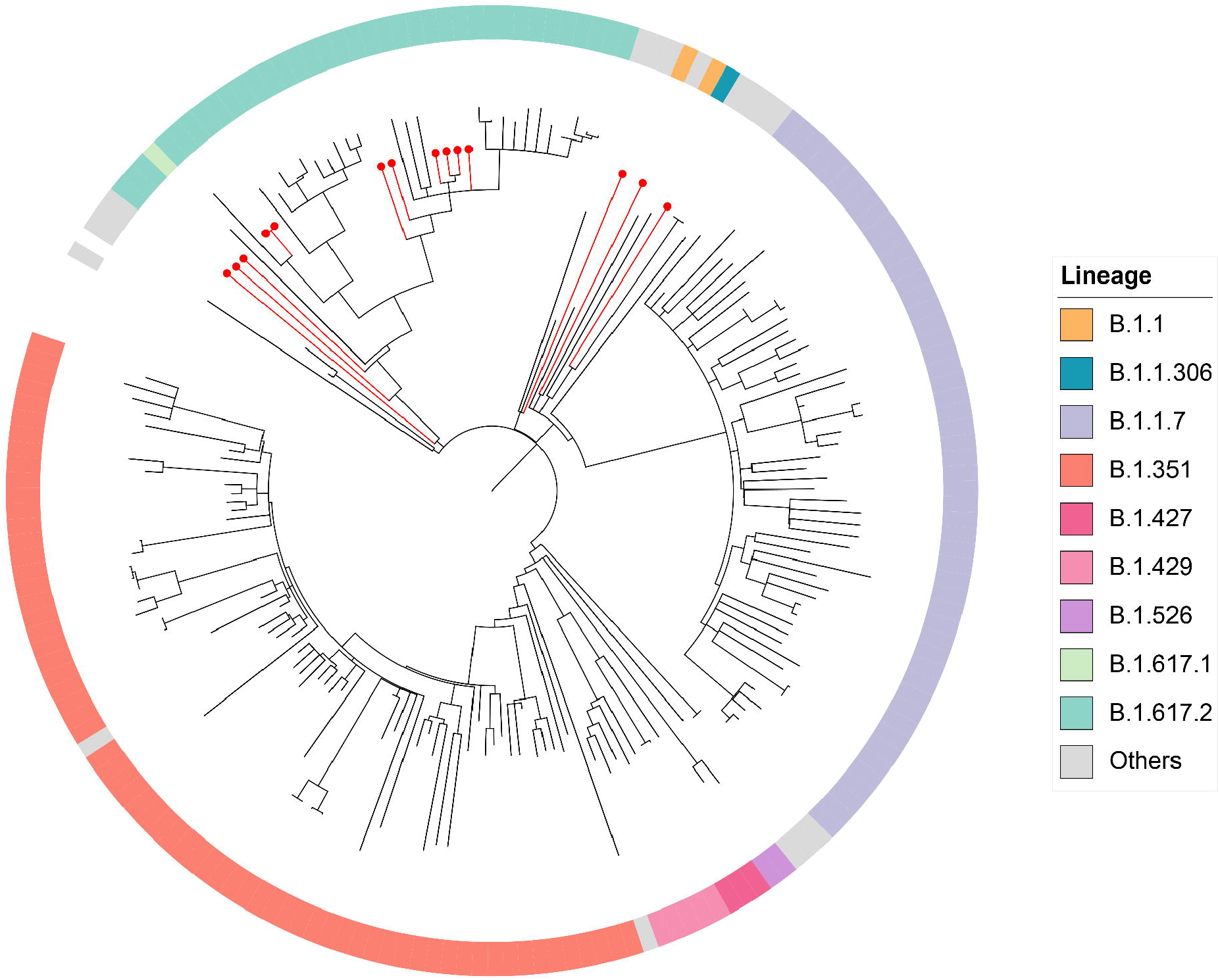
Phylogenetic analysis of Varanasi breakthrough samples with respect to global breakthrough samples. The Varanasi breakthrough samples were analyzed in the context of 172 (164 global and 8 Indian) SARS-CoV-2 genomes associated with breakthrough infections that were deposited in GISAID. The red lines indicate breakthrough samples from Varanasi. It appears that a high number of deposited samples belong to current VOCs/VOIs such as B.1.1.7 (Alpha), B.1.351 (Beta), B.1.617.2 (Delta) and B.1.427/B.1.429 (Epsilon)

## Data Availability

All sequenced samples were deposited on GISAID. Accession IDs for submitted samples are available in Supplementary Table S1. Accession IDs for samples not sequenced in this study and used in data analysis are available in Supplementary Table S1.

## Acknowledgements

We thank Tulasi Nagabandi for all the help and support with sequencing.

